# Impact of non-invasive ventilation on exacerbation frequency in COPD patients

**DOI:** 10.1101/2023.11.13.23298441

**Authors:** Maximilian Zimmermann, Georgi Margalitadze, Doreen Kroppen, Wolfram Windisch, Daniel S Majorski, Melanie Berger, Sarah B Stanzel, Maximilian Wollsching-Strobel

## Abstract

**BACKGROUND:** Acute exacerbations of COPD are key events in the natural course of the patients illness, as they significantly impair the health condition, accelerate the deterioration of lung function, worsen the prognosis for the patient and account for the majority of the COPD-related healthcare costs. Particularly in patients with a pre-existing non-invasive ventilation (NIV) therapy, a reduction of exacerbation frequency is crucial, as they are at high risk for a lasting morbidity and increased mortality.

**METHODOLOGY:** A prospective cohort study was conducted. Data from adult patients with the diagnosis COPD and existing High-Intensity NIV (HINIV) therapy from August 2021 to September 2023 was analyzed. Exacerbation history of moderate and severe exacerbations of the past 12 months and blood gases at initiation and during HINIV therapy were analyzed.

**RESULTS:** 20 patients were included (mean age 69.2±9 years; 70% female). After initiation of HINIV therapy the frequency of exacerbation displayed a significant reduction from 1.5±0.9 to 0.5±0.5 per year (p<0.001). In addition, improvements in pCO_2_ (73.0±22.0 mmHg vs. 44.0±4.8 mmHg; p<0.001), the pH (7.33±0.1 vs. 7.42±0.0; p<0.001) and HCO_3_^-^ (33.0±4.9 mmol/l vs. 27.9±3.2 mmol/l; p<0.001) were successfully demonstrated.

**DISCUSSION:** The present study demonstrates the positive effects of high-intensity NIV on exacerbation rate, measured by the number of moderate and severe exacerbations in one year. Most significant effects were observed when patients had a high number of exacerbations before the initiation of NIV therapy.

**What are the main findings?:**

- A significant reduction in the number of exacerbations was demonstrated with HINIV.
- There was a strong reduction in the frequency of exacerbations, especially in patients with a high frequency of exacerbations before the HINIV therapy was initiated.

**What are the implications of the main findings?:**

- The exacerbation history should be a more important factor in determining the indication for NIV therapy.
- Further studies on the pathophysiological causes must follow in order to comprehensively assess the therapeutic effects of NIV.

## Background

COPD is currently the fourth leading cause of death with approximately 3,2 million deaths in 2019 (1,2). In the course of the disease chronic hypercapnic respiratory failure (CHRF) can occur due to the failure of respiratory pump (3). COPD patients with CHRF have higher rates of unplanned hospital admissions and rapid clinical deterioration after hospitalization due to a severe exacerbation. Additionally, chronic hypercapnia is a decisive factor contributing to mortality (4,5). Therefore, reducing factors, influencing mortality, such as exacerbation rate, is an essential therapeutic goal. Non-invasive ventilation (NIV) with a facemask is the preferred choice of treatment to reduce mortality and hospitalization rate after acute exacerbation with persistent hypercapnia (6,7). However, these positive effects are evident only when NIV is used with the target of maximal possible pCO_2_ reduction (8,9). This ventilatory strategy, based on maximal relief of the respiratory pump is referred to as high-intensity NIV therapy (HINIV). The required inspiratory positive airway pressures (IPAP) vary individually, as they depend on side effects of the NIV therapy, patient tolerance and adherence to therapy.

Despite the central therapeutic goal of reducing the exacerbation rate in the treatment of COPD patients, given its significant impact on the health condition, acceleration of lung function deterioration, worsening prognosis of the patient and responsibility for the majority of COPD-associated healthcare costs, scientific data on the effects of a HINIV therapy on the exacerbation rate is insufficient. A large German study by Koehnlein et al. in 2014 was the first to observe reduction in mortality with the use of HINIV in COPD patients, but no conclusion could be drawn regarding the influence of HINIV on exacerbation frequency, as it was not part of the study (7). In a large British study investigating the effect of HINIV on hospitalization-free survival after acute exacerbation of COPD and acute respiratory failure, only severe exacerbations leading to hospitalization were recorded (6). The impact of HINIV on moderate exacerbation, however, is scientifically insufficiently documented (10).

The present analysis has addressed the lack of scientific evidence by initiating a study, evaluating the course of exacerbation retrospectively as well as prospectively, which compares the exacerbation frequency before initiation of HINIV and the exacerbation frequency after at least 12 months of HINIV therapy.

## Methodology

The study protocol was approved by the ethics committee of University Witten/Herdecke and was conducted in accordance to the ethical guidelines of the declaration of Helsinki (last revised in October 2013). Informed written consent was obtained from all patients.

### Patients

The data presented in this analysis are a preliminary analysis of the study, which was registered at the German register for studies (DRKS00029273) and investigates supplementary telemonitoring of COPD patients after experiencing a severe exacerbation. Data from adult patients with the diagnosis of COPD receiving HINIV due to hypercapnic respiratory failure (PrismaVent Type 30 (n=16) and type 40 (n=4), Löwenstein medical SE & Co. KG Bad Ems, Germany) between August 2021 and September 2023 were enrolled. Patients with mental retardation or those receiving invasive mechanical ventilation were not included in the study. The existing dataset of the main study was analyzed to determine availability of datasets regarding history of exacerbation as well as blood gases at the time of initiation or under existing HINIV therapy. The history of exacerbations is based on data from the clinical information system as well as from the anamnestic information provided by the patient, thus including exacerbations without hospitalization. Exacerbations that could be classified as moderate or severe according to the recommendations of the recent GOLD report were assessed (10). Therefore, exacerbations requiring treatment with a fast acting bronchodilator and oral corticosteroid ± antibiotics or requiring hospitalization were included.

### Study design and measurements

Demographic data (age, sex, exacerbation history) was evaluated for all patients. Additionally, laboratory parameters (pH, pCO_2_, pO_2_, HCO_3_^-^) were documented pre-therapeutically and after a minimum of 12 months after initiation of NIV therapy.

### Statistics

The data analysis was descriptive. Group comparison was conducted using t-test for normally distributed data and the Mann-Whitney-U testfor data with non-normally distributed data. Statistical significance was considered at a p-value of ≤0.05.

## Results

A total of 20 patients were included in the study (Figure 1). From those, 14 patients were female (70%). Average age of the patients was 69.2 ± 9 years. The included patients had been treated with NIV therapy for 4.9 ± 3.5 years. Long-term NIV therapy was initiated for 9 patients, as a result of acute NIV therapy with respiratory acidosis. Patients were using the HINIV therapy 8.9 ± 4.5h per day. The ventilation parameters from the time of blood gas analysis during therapy are listed in table 1. Significant improvements in blood gas analysis were observed during therapy, compared to the results before HINV initiation. Results of the blood gas analyses are detailed in table 2.

**Figure 1:**
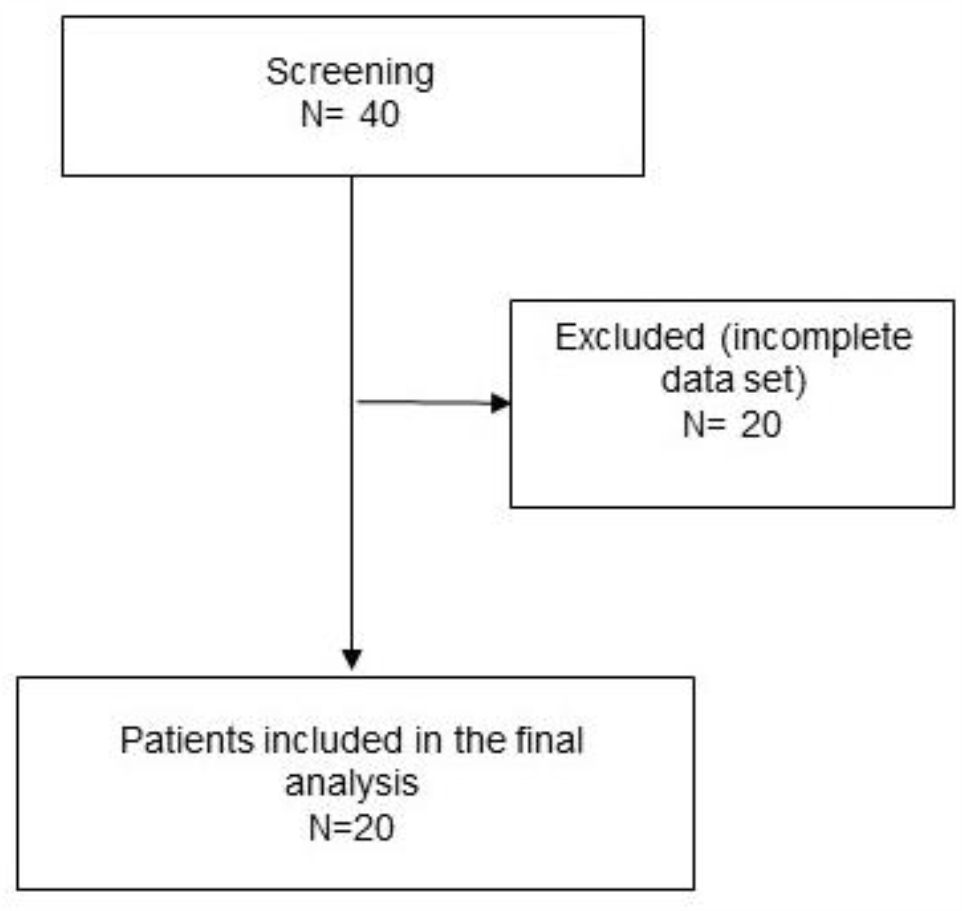
Diagram for patient inclusion

**Table 1:**
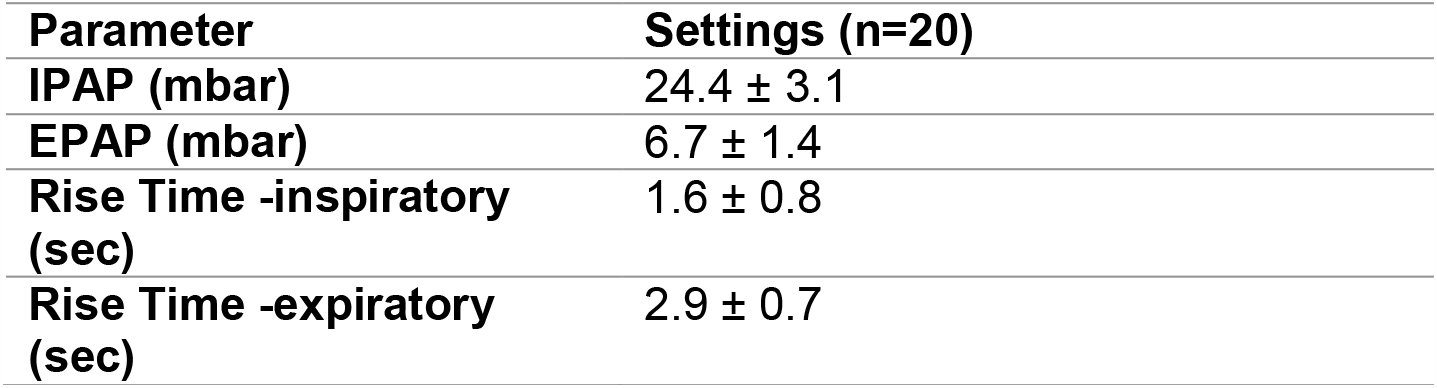
Ventilation settings during HINIV. Inspiratory positive airway pressure (IPAP); Exspiratory positive airway pressure (EPAP)

**Table 2:** Comparison of blood gas values before and after initiation of HINIV therapy of the total population (n=20)

|  | **Pretherapeuthic** | **HINIV** | **P- Value** |
| --- | --- | --- | --- |
| **pH** | 7.33 ± 0.1 | 7.42 ± 0.0 | <0.001 |
| **pCO<sub>2</sub> (mmHg)** | 73.0 ± 22.0 | 44.0 ± 4.8 | <0.001 |
| **pO<sub>2</sub> (mmHg)** | 79.0 ± 27.0 | 69.8 ± 17,3 | 0.2 |
| **HCO<sub>3</sub><sup>-</sup> (mmol/l)** | 33.0 ± 4.9 | 27.9 ± 3.2 | <0.001 |

A separate analysis was conducted for patients, who were initiated on a NIV therapy under stable conditions, without acute respiratory insufficiency. The results of this subgroup of 11 patients are presented in table 3.

**Table 3:** Comparison of blood gas values before and after initiation of HINIV therapy in patients, which had no acute respiratory insufficiency at the time of initiation (n=11)

|  | **Pretherapeuthic** | **HINIV** | **P- Value** |
| --- | --- | --- | --- |
| **pH** | 7.4 ± 0 | 7.4 ± 0 | 0.03 |
| **pCO<sub>2</sub> (mmHg)** | 61.4 ± 10.6 | 45.7 ± 5.3 | <0.001 |
| **pO<sub>2</sub> (mmHg)** | 84.6 ± 23,1 | 66.3 ± 15.4 | 0.08 |
| **HCO<sub>3</sub><sup>-</sup> (mmol/l)** | 32.9 ± 3.6 | 28.5 ± 3.9 | 0.01 |

Prior to the start of HINIV therapy, patients reported an exacerbation frequency of 1.5 ± 0.9 exacerbations per year. With HINIV therapy, this frequency was successfully reduced to 0.5 ± 0.5 per year (p<0.001). Individual differences were observed, as shown in Figure 2.

**Figure 2:**
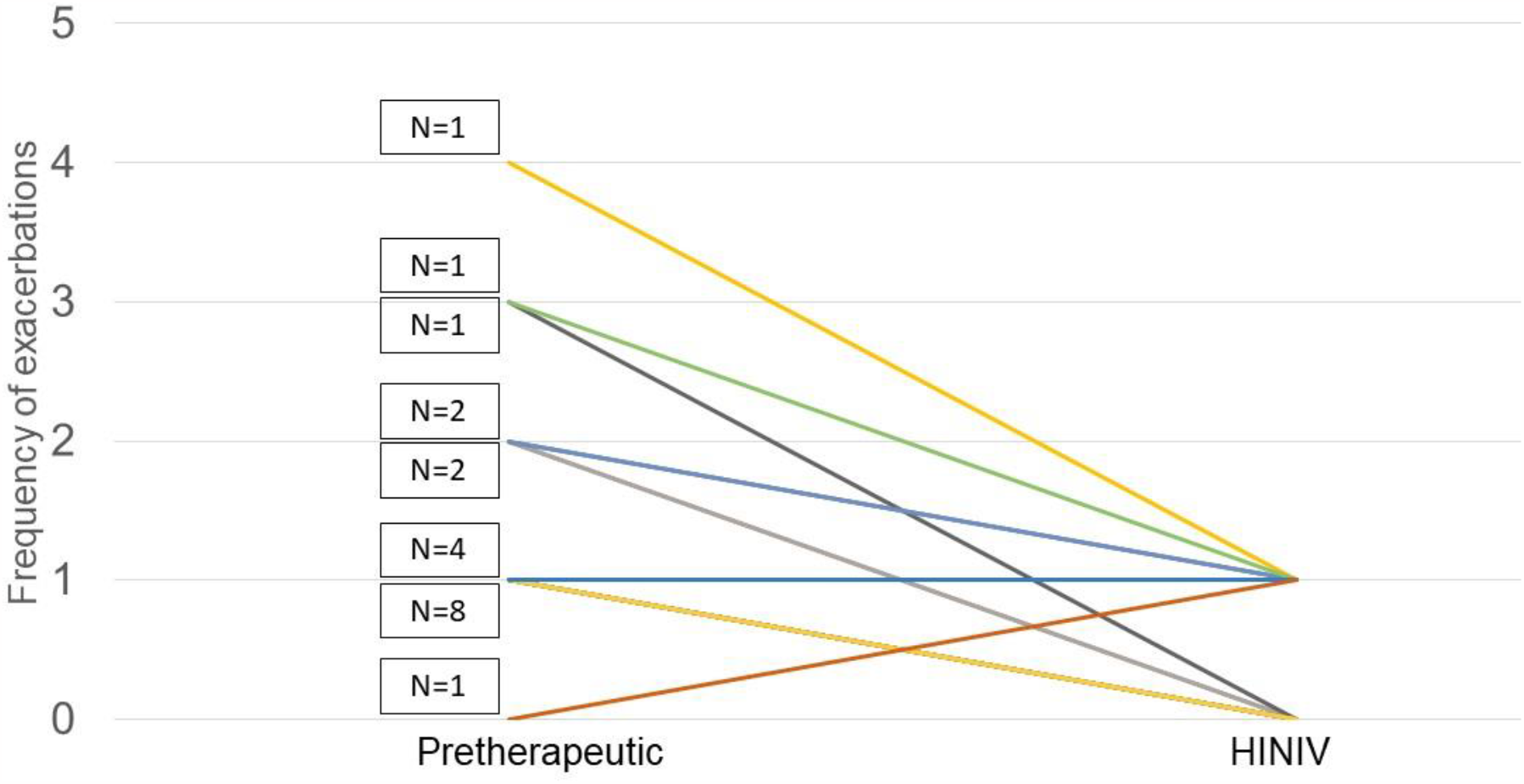
Illustration of individual differences in exacerbation frequency pre-therapeutic and after established HINIV therapy (N=20).

## Discussion

This study provides important information on reducing the frequency of moderate and severe exacerbations with established HINIV therapy. The following are the main findings, which will be discussed further:

Firstly, a significant reduction in the number of exacerbations was demonstrated with HINIV. Secondly, it has been shown that a significant reduction of hypercapnia is observed during HINIV therapy, both in the setting of acute respiratory failure and chronic respiratory failure. Thirdly, further blood gas analysis parameters showed a significant improvement under the established HINIV therapy. Fourthly, there was a strong reduction in the frequency of exacerbations, especially in patients with a high frequency of exacerbations before the HINIV therapy was initiated.

The significant reduction in exacerbation frequency under HINIV therapy differs meaningfully from previous studies that examined exacerbation frequency in hypercapnic COPD patients on NIV, but used significantly lower pressures and therefore could not reduce pCO_2_ (low intensity NIV) (11,12). This suggests that the impact of the NIV therapy on the exacerbation frequency can only be achieved with a sufficient augmentation of ventilation and thereby a significant reduction of hypercapnia. In patients with persistent hypercapnic respiratory insufficiency, a reduction in readmissions or mortality, following acute respiratory insufficiency, could be demonstrated after initiation of HINIV(6).

Furthermore, a recently published study by Hedsund et al. demonstrated, that the time to readmission with an acute respiratory insufficiency, due to an exacerbation within 12 months, could be reduced by HINIV, although, a statistical significance could not be shown due to insufficient recruitment (13). Nevertheless, a significant reduction in the number of exacerbations was observed, particularly in patients with frequent acute respiratory insufficiency. The lesser effects in comparison to the present study could be attributed to the fact, that normocapnic patients were included as well and no re-evaluation according to current recommendations was performed (14). All these studies only recorded the number of severe exacerbations that were hospitalized and did not include exacerbations that were treated in the outpatient setting.

The underlying pathophysiological mechanism of these reduced exacerbation rates resulting from HINIV therapy remains unknown. Besides the decrease in pCO_2_ levels, mechanical bronchial dilatation itself may alter the microbiological milieu. This has already been demonstrated with pharmacological bronchodilation (15–17). It has been shown that cytokines of the airways, which are elevated in patients with COPD, can be influenced by an effective treatment with bronchodilators (18). Whether these biomarkers can also be influenced by HINV therapy is the subject of ongoing research.

The present study has several limitations that should be taken into account. The data on exacerbations were mainly based on anamnestic information from the patient. Therefore, a higher incidence of exacerbations cannot be excluded. In addition, mild exacerbations were not recorded, so it is not possible to make any statements about the effect of HINIV on these exacerbations. The data was supplemented with information from the clinical information system. In addition, the number of participants is limited, resulting in a descriptive nature of data analysis, requiring larger subsequent studies are needed to validate the findings presented here.

## Summary

This study shows significant positive effects of HINIV on exacerbation rate, measured by frequency of moderate and severe exacerbations in patients with severe COPD. Greatest effects are observed when patients present with a high number of exacerbations prior to initiation of NIV therapy. To verify these effects in a larger collective, further studies need to follow.

## Data Availability

The authors intend to share individual deidentified participant data with any other unfunded or funded research-related purpose under appropriate circumstances. Please contact the corresponding author for more information.

## Abbreviations

CHRF: chronic hypercapnic respiratory failure
EPAP: Exspiratory positive airway pressure
HINIV: high-intensity NIV therapy
IPAP: Inspiratory positive airway pressure
NIV: Non-invasive ventilation

## Conflicts of interest

GM, MB: no conflict of interests. MZ, MWS, DK, DSM, SBS received travel grants from companies dealing with mechanical ventilation products. DSM reports open research grant from Philips Respironics/USA. MZ, WW received speaking fees from companies dealing with mechanical ventilation products. The study group received open research grants from Weinmann/Germany, Vivisol/Germany, Löwenstein Medical/Germany, VitalAire/Germany, and Philips Respironics/USA. The authors report no other conflicts of interest in this work.

## Funding

The main study was financially supported by Löwenstein medical SE & Co. KG (Bad Ems, Germany) with an open research grant without influence on the study design. This additional detailed analysis was performed without separate financial support.

## Author Contributions

All authors made substantial contributions to conception and design, acquisition of data, or analysis and interpretation of data; took part in drafting the article or revising it critically for important intellectual content; agreed to submit to the current journal; gave final approval of the version to be published; and agree to be accountable for all aspects of the work.

## Acknowledgments

This work has already been published as a pre-print under medrxiv.org.

## Notes

### Funding Statement

The main study was financially supported by Loewenstein medical SE & Co. KG (Bad Ems, Germany) with an open research grant without influence on the study design. This additional detailed analysis was performed without separate financial support.

